# Risk factors and symptom clusters for Long Covid: analysis of United Kingdom symptom tracker app data

**DOI:** 10.1101/2022.11.14.22282285

**Authors:** Elizabeth Ford, Harley Parfitt, Ian McCheyne, István Z. Kiss, Ruth Sellers

## Abstract

**Background:** Long Covid, characterised by symptoms after Covid-19 infection which persist for longer than 12 weeks, is becoming an important societal and economic problem. As Long Covid was novel in 2020, there has been debate regarding its aetiology and whether it is one, or multiple, syndromes. This study assessed risk factors associated with Long Covid and examined symptom clusters that might indicate sub-types.

**Methods:** 4,040 participants reporting for >4 months in the Covid Symptom Study App were included. Multivariate logistic regression was undertaken to identify risk factors associated with Long Covid. Cluster analysis (K-modes and hierarchical agglomerative clustering) and factor analysis were undertaken to investigate symptom clusters.

**Results:** Long Covid affected 13.6% of participants. Significant risk factors included being female (*P* < 0.01), pre-existing poor health (*P* < 0.01), and worse symptoms in the initial illness. A model incorporating sociodemographics, comorbidities, and health status predicted Long Covid with an accuracy (AUROC) of 76%. The three clustering approaches gave rise to different sets of clusters with no consistent pattern across methods.

**Conclusions:** Our model of risk factors may help clinicians predict patients at higher risk of Long Covid, so these patients can rest more, receive treatments, or enter clinical trials; reducing the burden of this long-term and debilitating condition. No consistent subtypes were identified.

## Introduction

A novel respiratory disease, now named Covid-19, emerged in China in late 2019, caused by a novel coronavirus SARS-CoV-2. By March 2020 the disease had spread around the globe and was named a global pandemic by the World Health Organisation on 11^th^ March 2020. Throughout the pandemic, hospitalisations and mortality have been a main focus in quantifying the severity and burden of the disease. However, it is now clear that these two blunt metrics hide an outcome of the disease which is hard to capture and measure, that of prolonged but moderate (i.e., patients are in their own homes) illness, which is nevertheless debilitating and incapacitating. This syndrome has been termed by patients “Long Covid” [1].

Research during the pandemic has indicated that a subset of patients with Covid-19, who may not have been hospitalised in the acute phase of the illness, will go on to develop a post Covid-19 syndrome: “a long term state of chronic fatigue characterised by post-exertional neuroimmune exhaustion” [2]. Commonly experienced symptoms include fatigue, brain fog, breathlessness, cough, chest pain, headache, gastrointestinal symptoms and widespread musculoskeletal pain [3, 4] Some research has identified that Long Covid is a relapsing remitting illness in which patients feel like they have recovered and then go on to experience symptoms again [5, 6].

Much initial evidence about Long Covid presentation originally came from patient-led research [7], medical blogs[8], self-report in social media [9], and cases reported in the main stream media. Later medical reports indicated that Covid symptoms were common in the three months following hospital discharge [10] and a review of mainly hospitalised patients found a prevalence rate of 43% among followed-up patients [11]. Among patients who were never hospitalised the prevalence has been less clear, as definitions, duration and recruitment strategies vary between studies. Estimates initially were that even among non-hospitalised patients, 34-38% experienced long-term symptoms [11, 12]. Later estimates for people who get Long Covid are between 2.3% and 37% [13], with the range in values attributed to a range of definitions. The Office of National Statistics (ONS) in the United Kingdom estimated that 13.7% of those infected experience Long Covid [14] and as of the 6^th^October 2022, 2.3 million people in the UK were estimated to be suffering from Long Covid of a duration of 4 weeks or longer (3.5% of the population) [14]. As our knowledge and understanding of the disease is only just over two years old, it is not clear what the long-term trajectory of the disease will be, but recovery appears to plateau quite quickly. In the UK 1.8 million people are estimated to have symptoms for 12 weeks or longer, showing only a 28% recovery rate between 4 and 12 weeks [14].

Established risk factors associated with ongoing Covid-19 symptoms past the acute phase include: being female [15-20], increasing age [15], prior heart, lung disease and asthma [15], severity of initial infection [15] and poor existing health [21]. However, these risk profiles are not always linear and an extensive review of electronic health records in the UK found that socio-demographic factors moderated the effect of key comorbidities [21]. Furthermore, much of this research focused on the risk of Long Covid on time periods prior to 12 weeks after initial infection (up to 12 weeks, but with focus on 4 weeks [15] and up to 60 days [22]), which falls short of NICE’s cut-off for defining Long Covid as “signs and symptoms” past 12 weeks [23].

One promising avenue for examining long Covid symptoms is the UK-based ZOE Covid symptom tracker app [24]. Over 4 million individuals have downloaded the app and are encouraged to track their symptoms on a daily basis. This huge “citizen science” dataset has been pivotal in a number of important Covid related studies, such as identifying symptom clusters [25], the diagnostic value of different signs and symptoms such as loss of smell [24, 26] predictors of hospitalisation [27], and vaccine side effects and effectiveness [28]. The phenomenon of Long Covid has also been studied in this data, with an attempt to estimate prevalence [15]. Many early projects used data from when many Covid patients did not have access to reliable testing. Using data from later in 2020, after the advent of universal testing, but before widespread vaccination programmes, we aimed to explore the potential of the Covid Zoe app data to inform practitioners, such as GPs, about Long Covid. This resulted in 2 aims for the project:

1. To determine the risk factors associated with Long Covid, defined as ongoing symptom beyond 12 weeks, to inform clinicians about who is likely to be at risk.
2. To determine if there was reliable evidence of different sub-types of Long Covid from the symptom tracker app data which might have differing risk factors.

## Methods

### Data Source

The COVID Symptom Study smartphone-based app (previously known as COVID Symptom Tracker) was launched in the United Kingdom on March 24^th^, 2020. It was developed by Zoe Global, in collaboration with King’s College London and Massachusetts General Hospital [24]. Anyone using the app is prompted everyday asking them to log their symptoms, even if they are healthy.

This project used data from individual users that was: 1) entered on registration of the app (location, demographic and medical history data) and 2) given in daily entries by each individual on their daily symptoms related to Covid. These two sets of data about each user are linked using a unique user ID. Further linked sets of data relate to tests for Covid-19 and to vaccinations against Covid-19. Users can enter a date on which they took the test, and later indicate if the test result was positive, negative or unclear, they can also provide details on their vaccinations.

App data provides an exciting opportunity to rapidly explore the emerging disease of Covid 19 during the pandemic. However, subscribers to a mobile app will not form a representative sample of the whole population, and instead, engagement with the app is likely to favour younger, more educated, more wealthy, and more tech savvy individuals [29]. In addition, there is likely to be substantial attrition of users’ engagement over time, leading to missing data or self-censoring of data. Thus, it is difficult to estimate population prevalence or other fixed parameters of a disease in this kind of data, as for research use, it is important to select a sample whose reporting within the app is consistent over time, and these users may not represent the population as a whole.

Data were accessed using SQL queries through the SAIL databank [30]. Full metadata associated with the Covid-19 Symptom Tracker Dataset can be found on the Health Data Research Innovation Gateway [31].

### App Variables

On registration for the app, personal characteristics about an individual such as their sex, ethnicity, age, BMI, smoker and location are obtained. In addition, the app also asks for information about illnesses or treatments that a patient may have such as lung disease, heart disease, cancer and whether they are pregnant, as well as whether the app user is taking blood pressure medications or aspirin.

At each log in, the app asks if the user has symptoms, or “feels physically normal”. Where they indicated they are “feeling not quite right”, a predetermined set of 39 Covid symptoms is presented. Each symptom can be indicated with a yes or no response, while a few can be answered as no, mild or severe. For example, if asked whether they are suffering from fatigue they can answer no, mild or severe, whereas persistent cough can only be answered as yes or no.

### Sample selection

To be included in our study, participants had to have met the following criteria:

- Logged on at least 120 days in total and have tested positive for Covid-19 between the 1st July and the 11th December 2020 (this cut off was decided to avoid the impact of vaccination on the study; UK vaccination roll out started 8^th^ December).
- Have a Body Mass Index of between 15 and 55 and be aged at least 18 years.
- Logged at least every 7 days after a positive test and within 7 days of the positive test.
- Logged at least 25 times between 14 and 84 days before the positive Covid test (to establish baseline health).
- Logged at least 50% of days between 12 and 16 weeks after the positive Covid test.

The sampled data was assessed for selection bias against a larger sample of 6801 patients who were over 18 and had BMI >15 and <55, had tested positive between 1^st^ July 2020 and 1^st^ January 2021, and had logged at least 120 times. Comparisons were made using two tailed Mann Whitney U test (for continuous data) and two tailed proportions Z tests (for categorical/ proportions data).

### Definition of Long Covid

Using the ONS cut-off of 12 weeks post infection, Long Covid was defined as having a statistically greater proportion of negative health status (sum of Covid symptoms compared to prior to infection) as identified by a proportions Z test (*P* < 0.05), in the period from 12-15 weeks after a positive Covid test compared to the period 2-12 weeks before a positive Covid result.

### Data pre-processing and feature engineering

Each participant’s symptom scores were normalised to account for their baseline health (i.e., before testing positive for Covid-19). As such, the data set was transformed from reporting the presence or absence of symptoms to reporting whether participants were experiencing symptoms more often, or at an increased severity than prior to infection. The normalised symptom scores were calculated, for each participant, by subtracting the average score, calculated from the period 12 weeks to two weeks before a positive Covid test, from the reported symptom score at every time point. Previous work using the same data source had neglected to account for baseline health status [15]. The baseline conditions were also used to establish a metric of a participant’s pre-existing health status

Following a similar approach to the work undertaken by KCL [15], symptom score metrics, based on the count of symptoms experienced at a higher rate than pre-Covid, over fortnightly periods, were created. Scores representing counts of all symptoms and those symptoms most heavily associated with Long Covid were created. All symptoms except fatigue, shortness of breath, diarrhoea and headaches had the same weighting (a score of one) in the calculation of the symptom score. Severe fatigue (struggling to get out of bed), significant shortness of breath (i.e., breathing is difficult even at rest), having diarrhoea five or more times a day, and having a headache all day, were considered twice as important in the calculation (i.e., a score of two).

For each consecutive two-week period after testing positive for Covid-19 (up until 8 weeks), it was calculated, for each symptom, whether the participant had experienced those symptoms at a greater frequency/ severity than during their period of baseline reporting.

A variable named “baseline health status” was created and was populated with a 1 if users indicated a status of “feeling not quite right” at least 10% of the time in the logging period before their first positive Covid test.

### Missing Data

It was deemed inappropriate, given Long Covid’s relapsing remitting nature [5] and high uncertainty over the reason for not reporting on any given day, to impute missing data about symptoms. For medical history variables, if a response was missing, it was assumed that the individual did not have that condition.

### Merged sources of data

A participant’s location, as identified by lower super output area (LSOA), was combined with relevant Index of Multiple Deprivation (IMD) data for England, Wales, Scotland, and Northern Ireland, available from their respective government websites [32] to give a proxy measure of relative socio-economic deprivation for each individual.

Postcodes were also combined with estimates of air pollution, a known risk factor for many cardiovascular and respiratory disease. Ambient background concentrations of nitrogen dioxide (NO_2_) and fine particulate matter (PM_2.5_), available at 1km^2^ resolution across the UK were taken from the UK-AIR resource [33] and combined with the participant’s location. It should be noted that the IMD and ambient air pollution level data are indicative neighbourhood level metrics and have been treated with caution.

### Data analysis

#### Aim 1: Which combination of risk factors best predicts Long Covid?

For univariate analysis of risk factors, two tailed Mann-Whitney U test was used to compare continuous data and two tailed Proportions Z test was used when dealing with count/categorical data.

To assess multiple predictors together, a logistic regression with LASSO penalisation model was chosen, due to its interpretability, its common use in epidemiological studies, including during the Covid-19 pandemic, [34-36] and its efficiency in training. Multi-variable models were run with two blocks; the first block contained medical history and demographic variables while the second included symptom scores in the 0-8 weeks following the positive Covid-19 result. This hierarchical design was chosen as early symptoms were highly predictive and other predictors in the model explained very little variance in the model if symptoms were added first.

Models were trained using balanced accuracy, not favouring recall over precision or vice versa. In the absence of an independent testing data set, 25% of the data were put aside for testing purposes. 20-fold cross validation was undertaken for validation purposes. Accuracy was assessed using Area Under the Receiving Operator Characteristic Curve (AUROC). Error analysis was undertaken on the final models to evaluate whether sub-sections of the sample were being correctly classified at lower rates than others.

#### Aim 2: Is there reliable evidence for sub-types of Long Covid?

There is reported great variability in the type and number of clusters identified from symptom clustering [37, 38] and so it was decided that three of the most common approaches should be taken forward for analysis: K-modes clustering [39], hierarchical agglomerative clustering [40] and factor analysis [41], to allow cross comparison between methods, thus allowing assessment of how robust any symptoms clusters are likely to be.

Clustering was only undertaken on the eleven most common symptoms of Long Covid to avoid a matrix that was too sparse. It was considered important that the symptom clustering report on any co-occurrence of symptoms, even if those symptoms were experienced regularly prior to getting Long Covid and as such, the data were not normalised by prior health status for the clustering analysis.

Both K-modes and HAC require the user to set the number of clusters and the optimal number of clusters was chosen by a combination of using ‘elbow plots’, which plot the decrease in mean square error with increasing number of clusters, and the silhouette score, which measures how similar an observation in one cluster is to points in adjacent clusters.

The Kaiser Meter Olkin test was carried to check whether the provided data was suitable for Factor analysis. A ‘middling’ score of between 0.7 and 0.8 was achieved indicating that factor analysis was a suitable statistical test to carry out [42]. Factor analysis used ordinary least squares to find the minimum residual solution. The number of factors was chosen by counting the number of eigenvalues in the correlation matrix over one, following the rule of thumb suggested by Kaiser [43]. To aide in interpretation of the factor matrices, they were rotated orthogonally using varimax rotation.

Clustering algorithms were run in python’s k-modes v0.11.0, factor_analyzer v.0.3.2 and sklearn v0.24.2.

## Results

### Sample Characteristics

After application of our eligibility criteria, our sample consisted of 4040 app users (see Figure 1). Of these users, 59.5% were female and 40.5% were male, they had a mean age of 54 years, and 97.5% were white. Participants were more likely to live in postcodes reflecting higher levels of income according to the Index of Multiple Deprivation (see Table 1).

**Figure 1:**
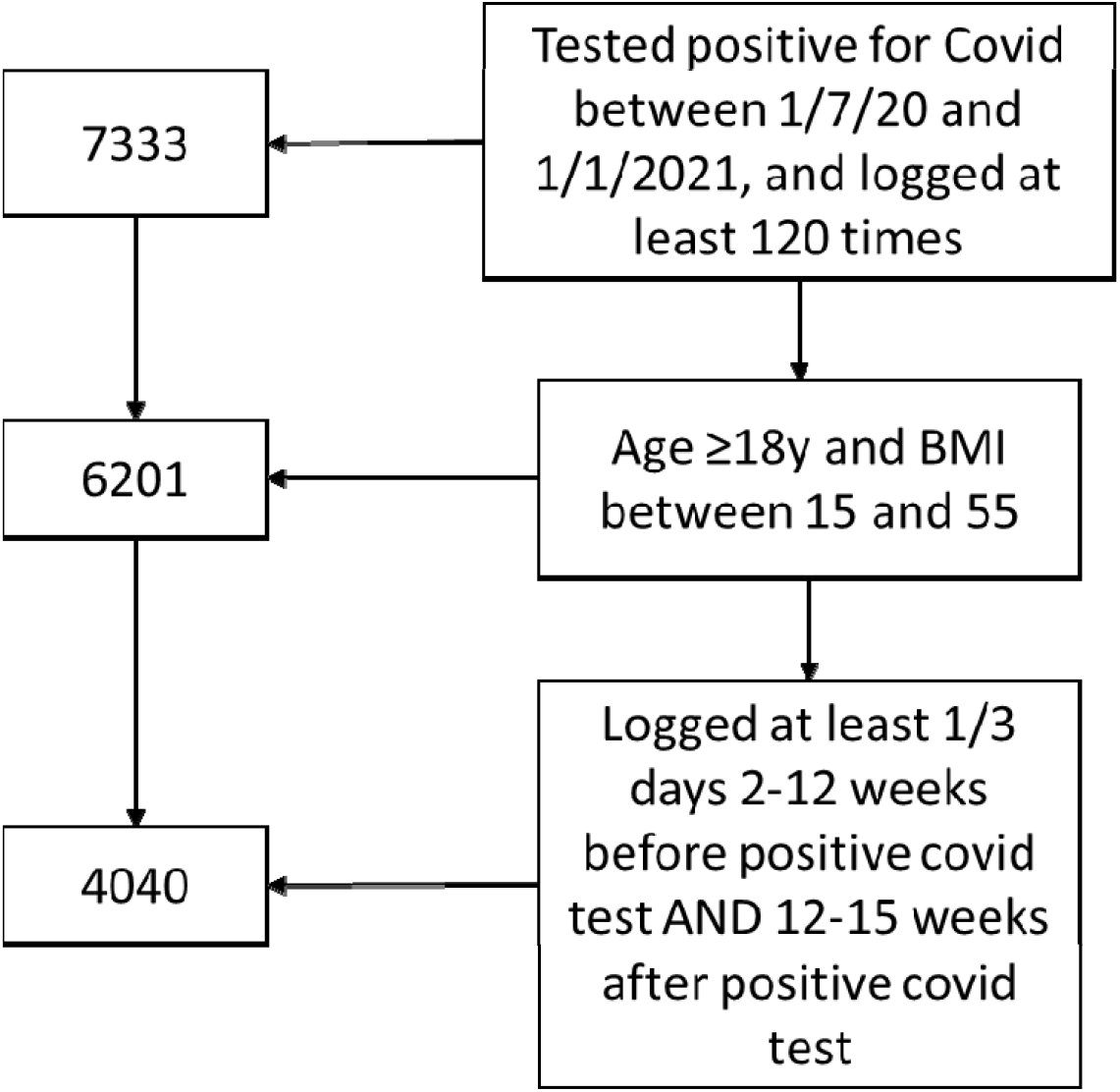
Sample selection process.

**Table 1:** Study participant characteristics from registration data.

| **Variable** | **Comparison Population** | **Sample (Including Long Covid)** |
| --- | --- | --- |
| Number | 6801 | 4040 |
| Body Mass Index (BMI) (mean) | 25.7 | 25.7 |
| Age (mean) | 52.3*** | 54.2 |
| Male (%) | 38.8* | 40.6 |
| Diabetes (%) | 4.2 | 4.3 |
| Lung Disease (%) | 12.5 | 12.3 |
| Heart Disease (%) | 3.5% | 3.7 |
| Kidney Disease (%) | 0.9 | 0.8 |
| Hay fever (%) | 38.5 | 37.3 |
| Asthma (%) | 12.3 | 11.5 |
| Cancer (%) | 1.9 | 2.1 |
| Limited Activity (%) | 8.8 | 8.5 |
| Smoker (%) | 3.3 | 3.3 |
| Is White (%) | 90.3*** | 97.1 |
| Total NO<sub>2</sub> (mean (IQR)) | 12.7 | 12.6 |
| Total PM<sub>10</sub>(mean (IQR)) | 8.7 | 8.7 |
| IMD decile (median, IQR) | 7 (5,9)* | 7 (5,9) |
| Does Not Take Vitamins (%) | 52.1 | 51.2 |
| Significant difference to sample at *0.1; **0.05; and ***0.01. Two tailed Mann-Whitney U test was used to compare continuous data and Two tailed Proportions Z test was used whe dealing with count data. |  |  |

We compared our selected sample (with BMI and age constraints and strict logging rules) against the reference population which included anyone who tested positive between 1^st^ July 2020 and 1^st^ January 2021 and who logged at least 120 times. We identified 6801 patients meeting these criteria. Applying our more stringent criteria for logging before, during and after Covid infection, we had a sample which was significantly whiter and more affluent that the reporting population, as well as slightly older with more males (shown in Table 1).

### Rates of Long Covid in Study Sample

13.6% of participants met the study criteria for ‘Long Covid’. Figure 2 provides a visual representation of Long Covid by plotting the cumulative normalised symptoms scores for the sample with and without Long Covid each day after a positive Covid result. 15.1% of the Long Covid group initially reported no symptoms at the time of their positive test. The graph shows that the Long Covid group had an initial resolution of symptoms up to 3-4 weeks after the positive test result and then very little improvement in symptom score after that point.

**Figure 2:**
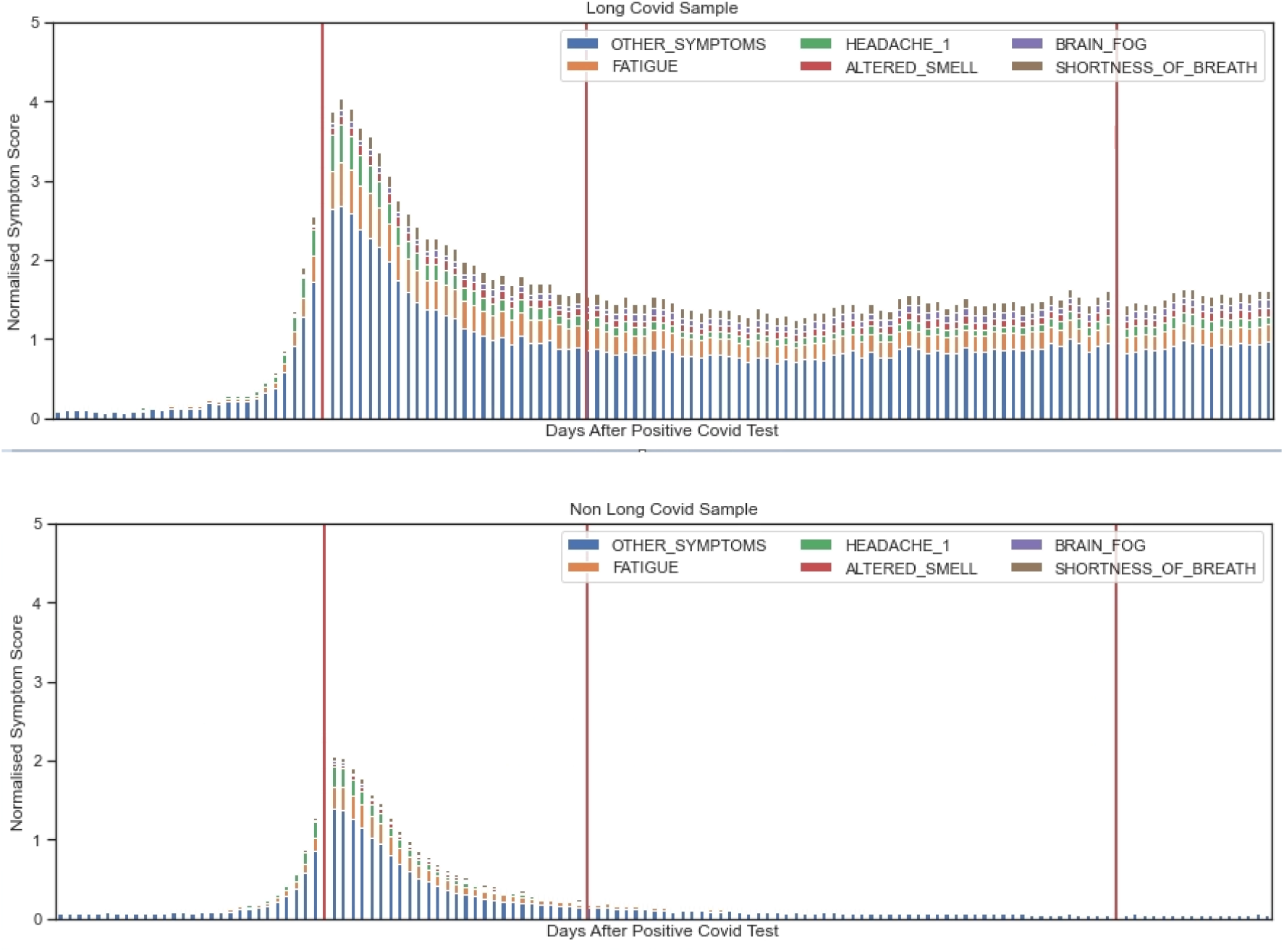
Visualisation of Cumulative Normalised Symptom Score. ***Top***: *Cumulative Normalized Symptom Score for sample with Long Covid;* ***Bottom***: *Cumulative Normalized Symptom Score for sample without Long Covid. Orange lines indicate date of test, 4 weeks after and 12 weeks after the test*.

### Aim 1: Risk Factors for Long Covid

In univariate analyses (shown in Table 2) Long Covid was significantly associated with being female, history of lung disease, hay-fever, asthma, previous limited activity, total NO_2_ and taking vitamin D or any vitamin supplement. The Long Covid group reported far higher rates of initial sickness, as well on-going negative health status. There were weak association between Long Covid and BMI and age (P <0.1).

**Table 2:** Characteristics of the sub samples with and without Long Covid.

| **Variable** | **Without Long Covid** | **With Long Covid** |
| --- | --- | --- |
| Number | 3489 | 551 |
| Body Mass Index (BMI) | 25.6 | 26.1* |
| Age (mean) | 54.0 | 55.5* |
| Age Group (18-49; 50-69; >70 (numbers) | 965, 2113, 411 | 141,339, 71 |
| Age Group (18-49; 50-69; >70) (%) | 27.7, 60.6, 11.8 | 25.6, 61.5, 12.9 |
| Male (%) | 41.9 | 32.1*** |
| Diabetes (%) | 4.4 | 4.0 |
| Lung Disease (%) | 10.9 | 15.4*** |
| Heart Disease (%) | 3.6 | 4.4 |
| Kidney Disease (%) | 0.8 | 0.9 |
| Hay fever (%) | 36.6 | 42.0** |
| Asthma (%) | 11.1 | 14.1** |
| Cancer (%) | 1.3 | 2.3 |
| Limited Activity (%) | 7.8 | 13.2*** |
| Smoker (%) | 1.5 | 1.9 |
| Is White (%) | 97.1 | 96.7 |
| Total NO2 (µg.m-3) (mean (IQR) | 11.1 | 12.4*** |
| Total PM2.5 (µg.m-3) (mean (IQR) | 8.7 | 8.6 |
| IMD decile (median, IQR) | 7 (4,9) | 7 (5,9) |
| Does Not Take Vitamins (%) | 52.4 | 42.6*** |
| Takes Vitamin D | 21.8% | 28.3*** |
| Negative Health Status (0-2 Weeks) (%) | 60.5 | 84.5*** |
| Negative Health Status (2-4 Weeks) (%) | 27.7 | 67.7*** |
| Negative Health Status (4-6 Weeks) (%) | 14.9 | 59.3*** |
| Negative Health Status (6-8 Weeks) (%) | 10.3 | 56.6*** |
| Maximum Number of Symptoms (median (IQR) | 1(1,3) | 3(1,4)*** |
| Ill Before (%) | 7.8% | 10.5%** |
| Asymptomatic (%) | 37.6% | 15.1%*** |

Significant difference to sample at \*0.1; \*\*0.05; and \*\*\*0.01.

Symptoms during the 0-8-week period following infection were highly predictive of Long Covid. From 4-6 weeks onwards, fatigue and olfactory issues became the stronger predictors of Long Covid, alongside number of symptoms and general health status as overall measures. The risk factors of medical history and demographics were moderated by age. Having a pre existing health condition was positively associated with Long Covid in all age groups up to 70 years but over 70 it was negatively associated. Being male was negatively associated up to 70 years but positively thereafter. Individual associations are shown graphically in supplementary materials.

### Multi-variable analysis to predict Long Covid

A hierarchical logistic regression with two blocks and LASSO penalisation was run using medical history, demographics (block 1) and early symptoms (block 2) as predictors.

### Block 1: medical history and demographics but no symptom data

This model achieved an AUROC of 0.69 (Figure 3). The retained features in this model were gender, baseline health status, limited activity, no vitamins, takes vitamin D. The strongest features in this model were being female (0.43), followed closely by having a condition that limited physical activity (0.42).

**Figure 3:**
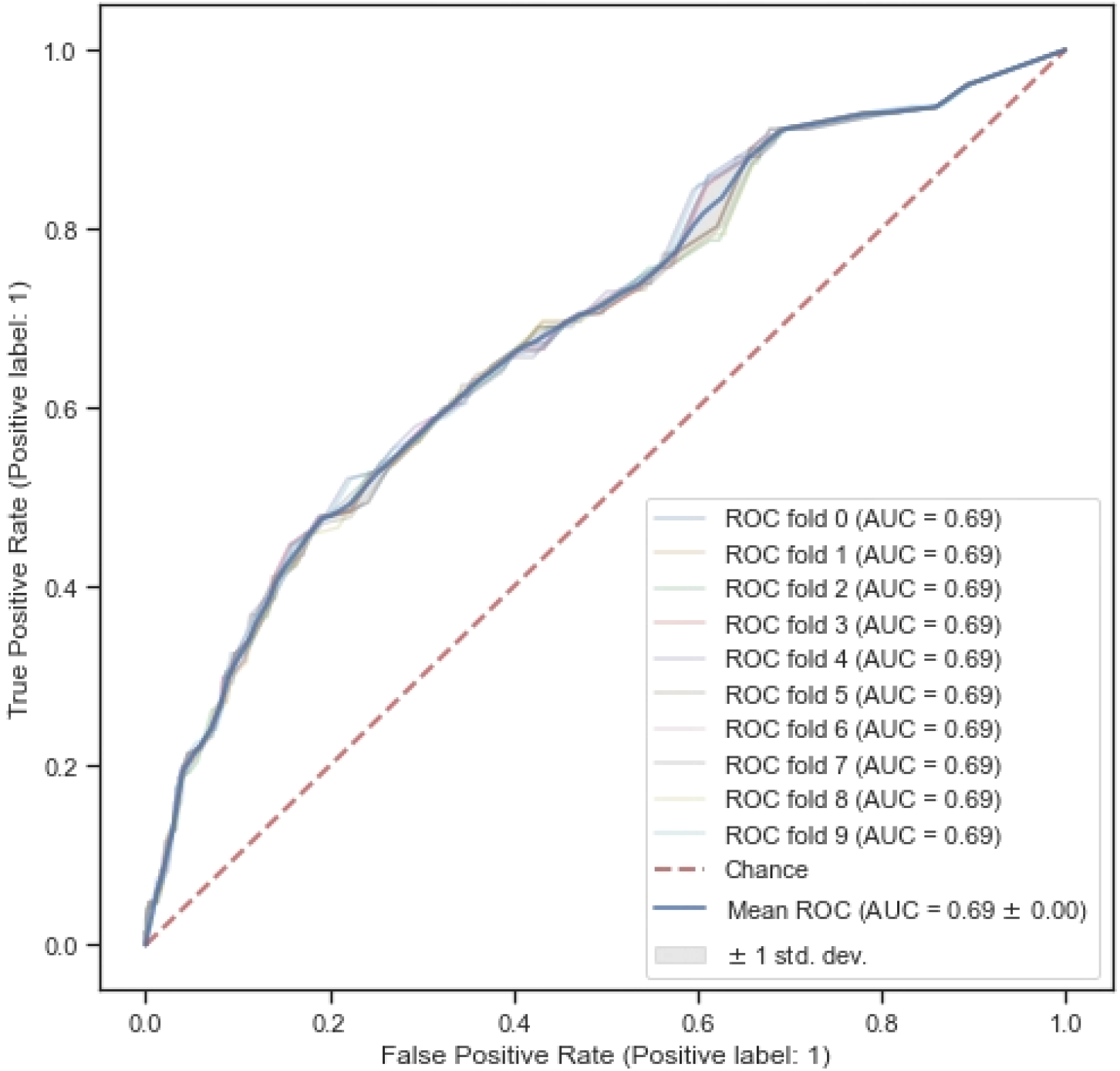
Receiver operating characteristic curve for the model with medical history and demographics.

Error analysis of the early-stage model found that of those participants who were defined as having Long Covid *and* exhibiting no initial symptoms, the model could only accurately predict 1/21 correctly (4.8%). This compared to 85 out of 120 (70.1%) with symptomatic initial infection.

### Block 2 with medical history, demographics and Covid symptom data up to 8 weeks

This model retained the following variables: gender, max symptom score in first two weeks, baseline health status, limited activity, no vitamins, takes vitamin D. The AUROC of the model was 0.77, thus the model was substantially improved by adding symptom data from the first two weeks of the illness (Figure 4). The symptom score in the first two weeks had a coefficient of 0.34.

**Figure 4:**
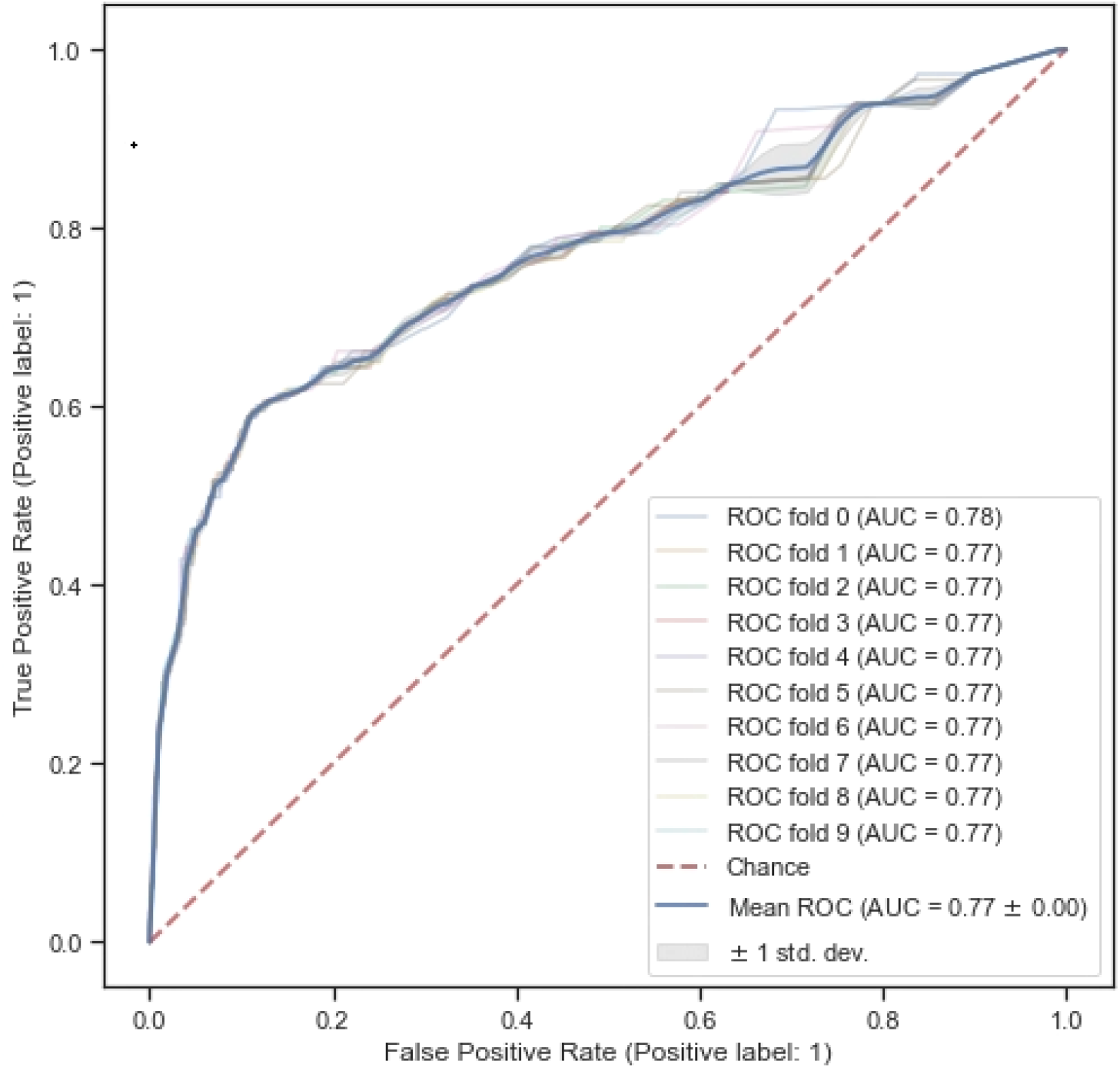
Receiver operating characteristic curve for the model with medical history, demographics and symptom data.

Error analysis of the reduced model found that of those participants who were defined as having Long Covid *and* exhibiting no initial symptoms, the model could only accurately predict 5/21 correctly (23.8%). This compared to 80 out of 120 (66.1%) with symptomatic initial infection. (Recall = 61.7%)

### Aim 2. Is there reliable evidence for sub-types of Long Covid?

The results of the K-modes clustering provided no strong evidence for symptom clusters. The Hierarchical Agglomerative Clustering and the Factor Analysis produced differing results.

### Hierarchical Agglomerative Clustering

This analysis showed some evidence for a 2-cluster fit from the silhouette score; a score of over 0.4 is acceptable. When cluster loadings were plotted, 2 clusters appeared to be a reasonable cut off (Figure 5). As Figure 6 shows, the two factors seemed to indicate mild and severe versions of Long Covid, as loading patterns were similar between the two factors, but loadings were much stronger in the second cluster.

**Figure 5:**
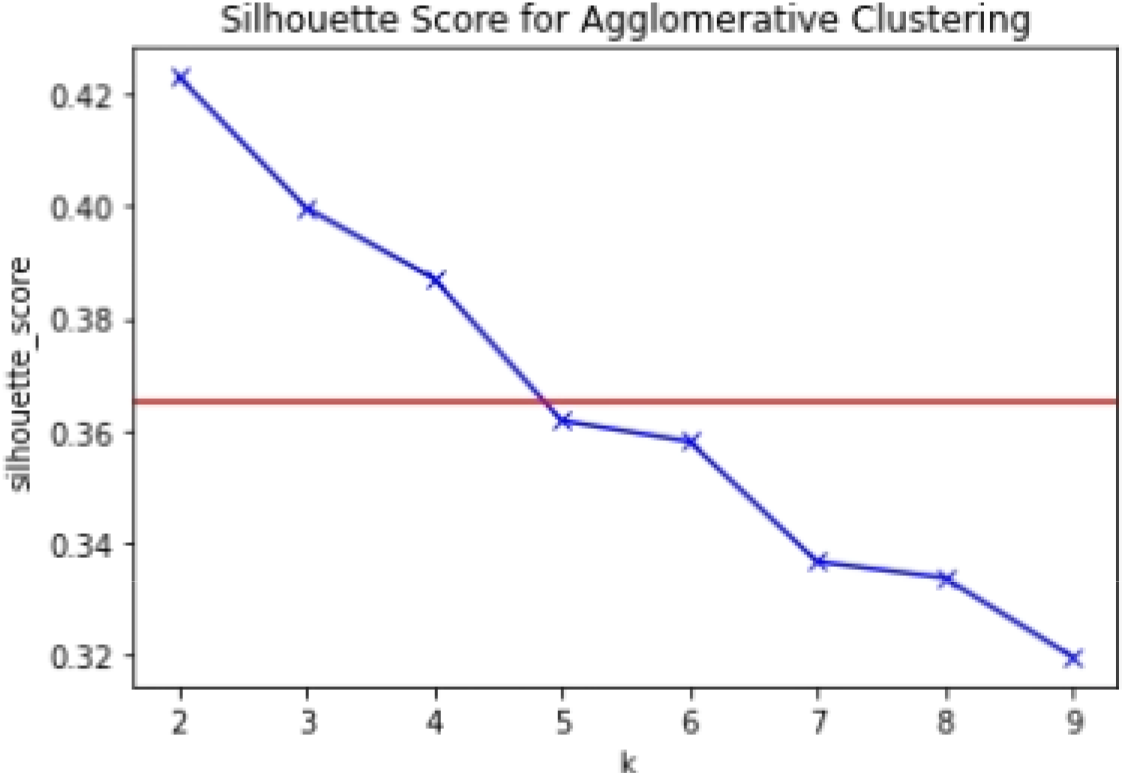
Plot of Silhouette Score for Hierarchical Agglomerative Clustering (>0.4 is an acceptable score).

**Figure 6:**
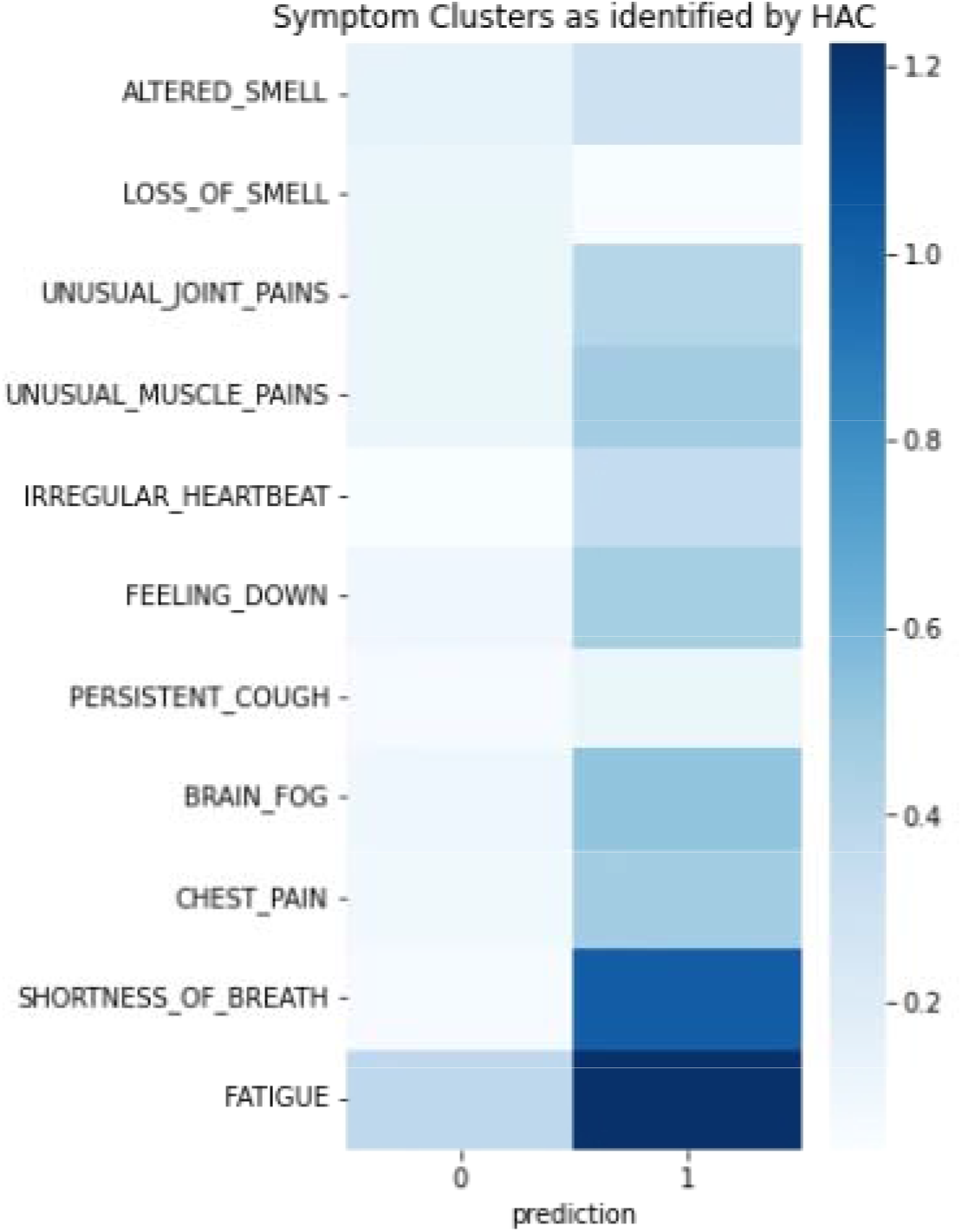
Cluster Loadings in the HAC 2-Cluster model.

### Factor Analysis

Factor analysis identified four potential factors, explaining up to 45% of the variance in co occurrence of symptoms, but these differed slightly by men and women as shown in Table 3.

**Table 3:**
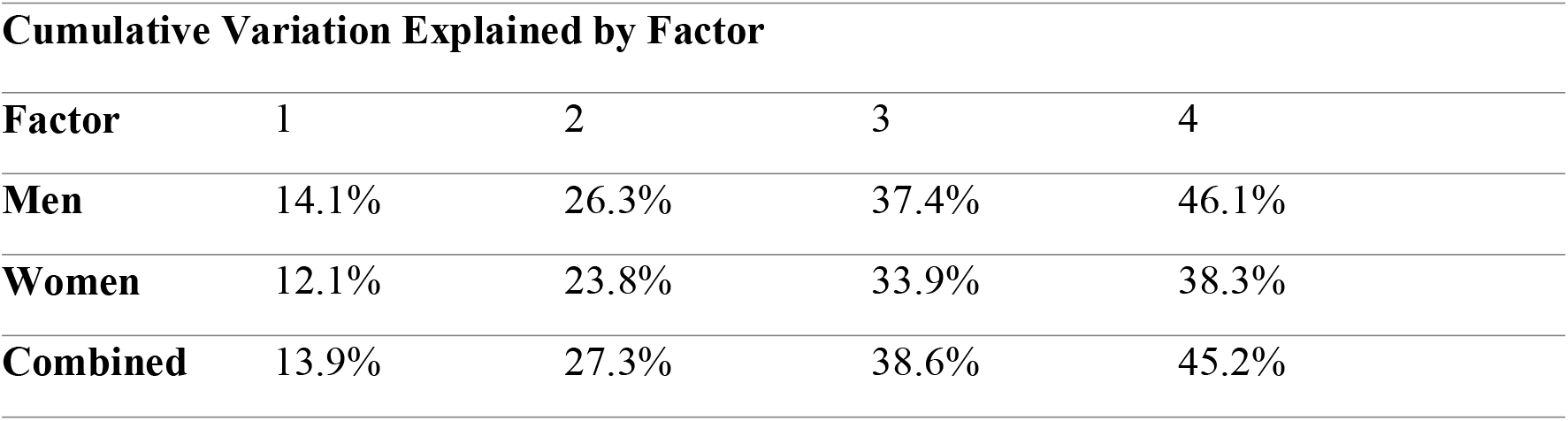
Cumulative Variation Explained by Factors.

| Cumulative Variation Explained by Factor |  |  |  |  |
| --- | --- | --- | --- | --- |
| Factor | 1 | 2 | 3 | 4 |
| Men | 14.1% | 26.3% | 37.4% | 46.1% |
| Women | 12.1% | 23.8% | 33.9% | 38.3% |
| Combined | 13.9% | 27.3% | 38.6% | 45.2% |

Examining Figure 7 (all participants), the first factor is characterised by extreme fatigue and predominantly neurological symptoms including brain fog and feeling down. The second factor is characterised by joint and muscle pain, with the third characterised by chest pain, shortness of breath, and irregular heartbeat, perhaps suggestive of long-term damage to the heart and lungs. The fourth factor is only visible when considering the male and female fourth factors in isolation, suggesting an impact on the olfactory system only.

**Figure 7:**
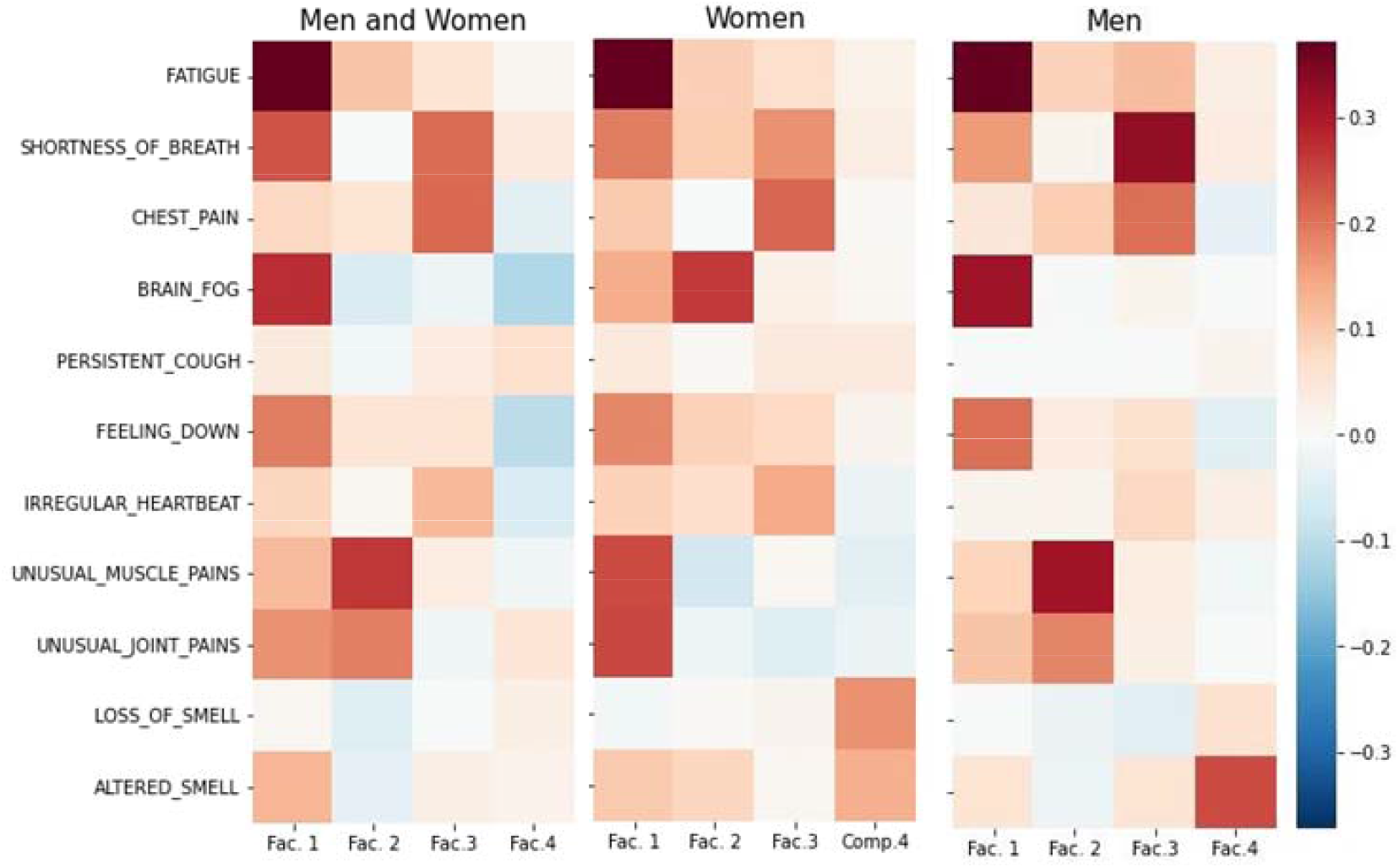
Factor Analysis heatmap showing loading of symptoms onto factors.

## Discussion

This study aimed to examine risk factors associated with Long Covid, to inform clinicians about who is likely to be at risk. We identified that being female, having limited activity and pre-existing health conditions before Covid, as well as having a higher range of symptoms at the onset of Covid illness, were at higher risk of having Long Covid at 12 weeks post infection. The range of variables associated with Long Covid looked different in the over 70s age group. Using the range of variables available in the symptom tracker app, we were able to predict Long Covid at the onset of illness with 69% accuracy, and after 8 weeks of symptoms with 77% accuracy, with the highest rate of error (false negatives) in people who had an asymptomatic Covid infection.

Our second aim was to examine whether there was reliable evidence of different sub-types of Long Covid. After trying three different clustering approaches, we found no consistent factor or cluster structure across methods. Factor analysis offered a solution with four factors although the differential between these was not very clinically meaningful, except for one factor which potentially pointed to lung and heart complications.

Being female was a significant risk factor, which supports the growing consensus in the academic literature [18-20, 44-46]. However, women had lesser risk than men in the >70 age cohort, which supports the conclusions of Sudre et al [44] and adds weight to the hypothesis that female sex hormones might, to a degree, be a factor in causing Long Covid [47]. However, these findings contrast with hospitalisation and mortality data where being male gives a high risk for severe disease and death [48-51]. Having poorer existing ill-health was a significant predictor for people up to 69 years, which has also been found as a significant predictor in other research [21]. Additionally, having limited activity prior to Covid was associated with Long Covid and might be a proxy for underlying chronic conditions which limit mobility or energy levels.

There was no evidence that smoking was associated with Long Covid. Interestingly, early in the pandemic, smokers were under-represented in Covid-19 mortality figures [52] and there are biologically plausible mechanisms that smoking might offer protection from Covid-19, and subsequent Long Covid. These include the reduction in ACE2 expression (a potential Sars-COV-2 cell receptor [53]), nicotine’s anti-inflammatory qualities and even potential immunity caused by the presence of heat shock proteins in the respiratory tract [52]. However, some subsequent research has found a positive link between smoking and risk of Long Covid [54, 55]. There was some suggestion of a negative association with diabetes in our data. There are reports that certain diabetes treatments (SGLT2 inhibitors and GLP1RA) could help treat Long Covid through improvements to blood pressure, inflammation and hyperglycaemia [56]. Similar results were also found with heart disease sufferers in the >50 age category suggesting commonly used medicines, such as statins, which also regulate ACE2 inhibition and immune inflammation [57] might help prevent and treat Long Covid.

### Strengths and limitations

The core strength of using app-based self-report data is that this data was collected daily, in real time, during the first 9 months of the Covid pandemic in the UK, and therefore has high reliability. Our sample is also reasonably large. Our study used a statistically robust way of defining Long Covid and the reported Long Covid rate was directly comparable to the ONS’ estimate (13.6% vs 13.7%) and falls within current range of estimates (2.3 %– 37%)[13]. However, we note a number of limitations with the data source, timing of the study, and potentially with our analytic approach.

The main limitation for study generalisability is the representativeness of the sample. Our sample had an over-representation of older, female, white and higher income individuals than would be expected from a representative sample in England. It is not fully understood how these selection biases would affect our estimates for Long Covid, its risk factors and underlying causes. Although limited in stratifying by age and gender, this research found that comorbidities were moderated by these characteristics, and this is supported by other research which looked at multiple sociodemographic groups [21]. We excluded children from the sample, so our model cannot predict Long Covid in these younger age groups, although it is clear that some children can experience Covid symptoms for >4 weeks [58, 59]. Long term Covid is still not well understood or researched in children, and this should be an avenue for future work.

The second data limitation is that of missing data. We tried to minimise the impact of missing data by selecting individuals who had at least 120 data entries and estimate symptom scores over fortnightly periods. There is some evidence around Long Covid being a relapsing – remitting illness, with periods of recovery followed by resurgence of symptoms [5, 7, 60]. Our study is underpinned by the assumption that individual reporting behaviour and the perception of symptoms did not change throughout the study period. For example, it was assumed that the likelihood of reporting a positive or negative health report remained constant over the study period. The plausibility of this assumption was not thoroughly investigated. In addition, we treated missing data on medical history variables as negative, which may have under-estimated prior medical conditions. Furthermore, the UK’s vaccination programme potentially introduced a source of bias, as individuals may have received the vaccine during their 12-week post-infection period, and this was not assessed in the study. Some individuals have reportedly shown an improvement in symptoms after the vaccine [61].

Our ability to study potential risk factors for Long Covid was limited by the questions asked in the app at registration, by the symptoms recorded in the app, and potentially by low base rates of some risk factors (e.g. smoking rates are low in England [62]). Understanding of the risk factors for and presentation of Covid has evolved rapidly over the last 20 months, and the app has been regularly updated to accommodate this. However, data entered by users in earlier periods of the pandemic will have been limited to the questions asked at the time. We note that all of our participants were pre-vaccine at the moment of their positive Covid test, and vaccine coverage is likely to have reduced the risk of Long Covid [63] in individuals vaccinated prior to getting infected, although it is not clear if it has changed the pattern of risk factors. We also did not cover periods of differing Covid variants, which may cause differing symptom patterns and risk profiles, so our study should be repeated in more recent data and examine the effects of vaccination and known dominant variants.

Lastly, we still have limited understanding of risk factors for Long Covid in those who test positive for Covid but remain asymptomatic. Our model was not able to predict Long Covid well in these cases, and they may have a different risk profile.

### Implications for care planning and management

These findings are of interest and contribute to clinical understanding of Long Covid. Patients with a severe set of symptoms in the acute phase of the illness will need to be followed up with extra support as they are likely to experience a longer duration of symptoms. Our findings accord with emerging evidence that females, older adults, individuals with pre-existing long term or mobility limiting conditions might be more likely to have a longer duration of symptoms. Clinicians caring for people after the acute phase of Covid, such as general practitioners, should be aware of these risk factors and plan their management of, and communication with, patients accordingly. Long Covid services have now been set up in most regions and a referral pathway for long term support, including respiratory physiotherapy, fatigue, neurological, and mental health management, should be available for these patients.

Our predictive model had an accuracy, measured by AUROC of 0.77, which is thought to be an acceptable standard [64]. However, before any implementation in clinical practice, developers of prediction models should give careful thought to the potential for false negative and false positive predictions, and whether patients could be harmed by these; the positive predictive value (PPV; the number of people predicted to get the condition who do, in fact, get the condition) of prediction models and diagnostic tests is related to sensitivity and specificity (as depicted by the ROC) <u>and</u> the prevalence of the condition in the target population. Even when an AUROC is fairly high, the PPV can still be low if the population prevalence is low, due to high numbers of false positives. Ethically, it is additionally imperative to have a well-resourced clinical pathway in place before any implementation of a risk calculator. Flag positive patients must be offered a service or intervention which meaningfully treats or supports their condition or reduces their risk, according to ethical screening principles [65].

## Conclusions

Using self-report app data collected during the pre-vaccine phase of the pandemic, we found that a combination of early symptoms, sex, baseline health status, and prior limited activity was able to predict long Covid in symptomatic individuals with good accuracy. Severity of symptoms in the first 8 weeks of illness was the strongest predictor of Long Covid. We did not find replicable clusters of symptoms suggesting more than one type of Long Covid among these patients, so cannot support hypotheses that there are measurable sub-types of Long Covid. These findings will be informative for practitioners caring for individuals with Covid and may help inform rehabilitation services.

## Data Availability

All data are held securely in the SAIL databank and are not openly accessible to other researchers, except with a project proposal and data access request made directly to SAIL.

https://saildatabank.com/

