## supplementary material for "Risk factors and symptom clusters for Long Covid: analysis of United Kingdom symptom tracker app data"

*Supplementary Materials: Risk Coefficients for Long Covid for Individual Features, by Age Groups*

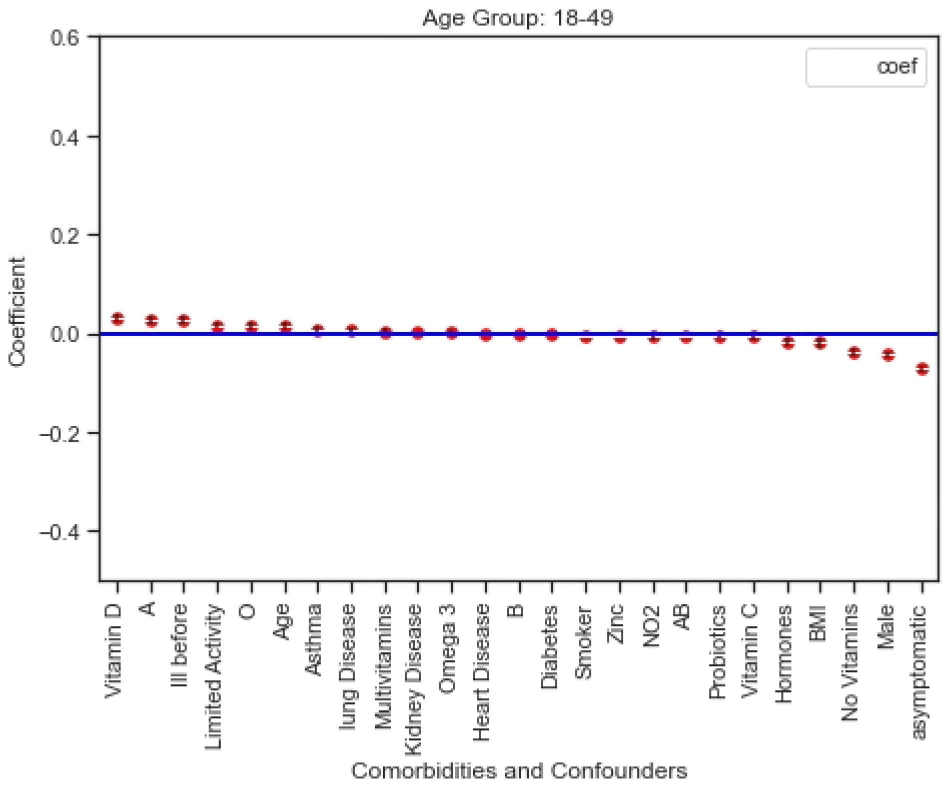


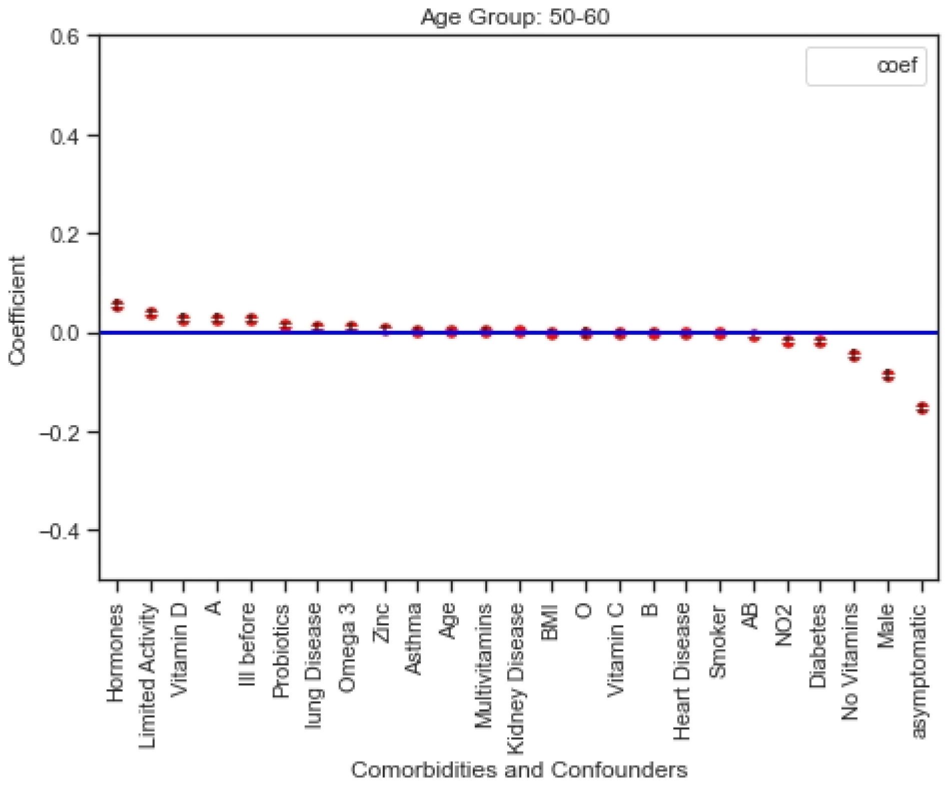

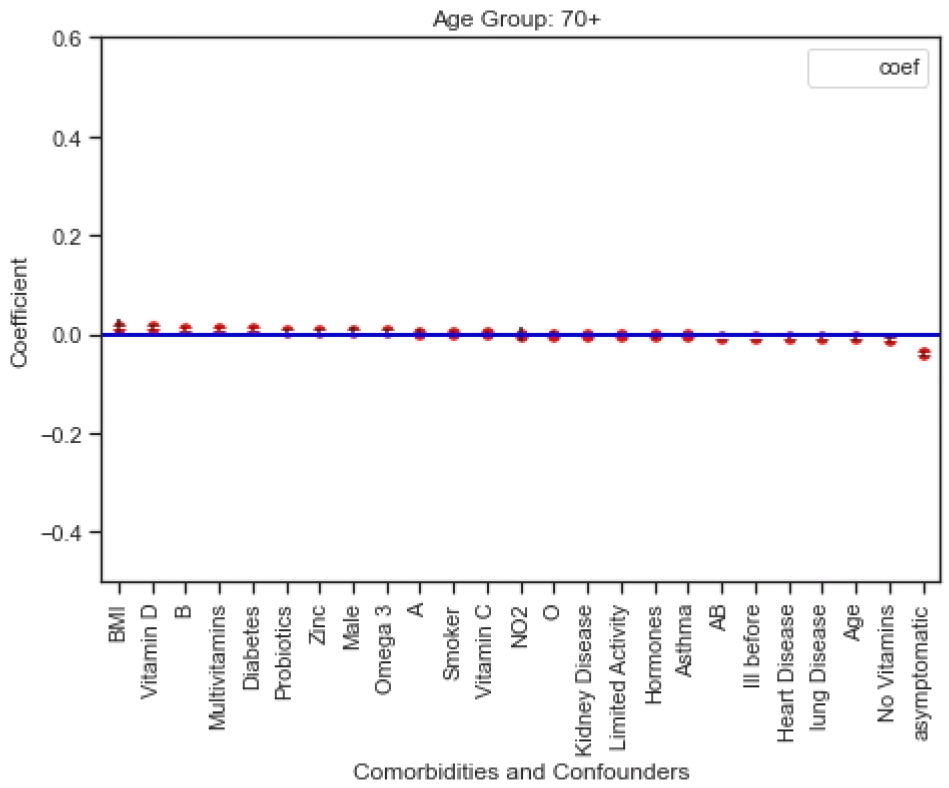
Age Group: 70+
coef
Coefficient
0.6
0.4
0.2
0.0
−0.2
−0.4
BMI
Vitamin D
B
Multivitamins
Diabetes
Probiotics
Zinc
Male
Omega 3
A
Smoker
Vitamin C
NO2
O
Kidney Disease
Limited Activity
Hormones
Asthma
AB
Ill before
Heart Disease
lung Disease
Age
No Vitamins
asymptomatic
Comorbidities and Confounders

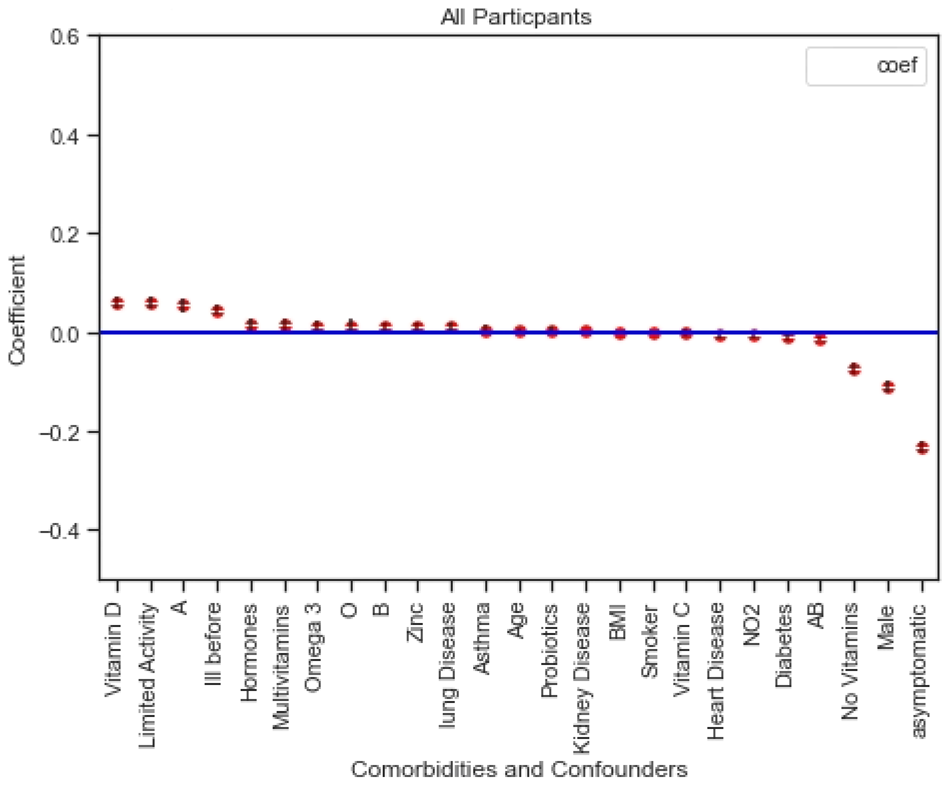
All Particpants
coef
Coefficient
0.6
0.4
0.2
0.0
−0.2
−0.4
Vitamin D
Limited Activity
A
Ill before
Hormones
Multivitamins
Omega 3
O
B
Zinc
lung Disease
Asthma
Age
Probiotics
Kidney Disease
BMI
Smoker
Vitamin C
Heart Disease
NO2
Diabetes
AB
No Vitamins
Male
asymptomatic
Comorbidities and Confounders
